# Assessment of Ontario-purchased commercially available milk products for the presence of influenza A viral RNA

**DOI:** 10.1101/2024.06.03.24308235

**Authors:** Juliette Blais-Savoie, Winfield Yim, Jonathon D. Kotwa, Lily Yip, Robert Kozak, Allison McGeer, Samira Mubareka

## Abstract

Given the widespread presence of influenza A(H5N1) viral RNA in pasteurized milk in the United States, we tested 117 samples of pasteurized cow’s milk purchased from retail outlets in Ontario, Canada in April 2024 for the presence of influenza A viral RNA. No influenza A viral RNA was detected.

## Introduction

Bovine infections with H5N1 highly pathogenic avian influenza (HPAI) virus were first identified in the United States in March of 2024. As of May 30, 2024, 66 herds of dairy cattle across 9 states have tested positive.^1^ In late April, multiple studies identified the presence of H5N1 HPAI viral RNA (vRNA) in commercially available milk across the United States, suggesting that infections in dairy cattle were much more widely disseminated than had been initially believed.^2^ Genomic analysis suggests the cattle outbreaks originated from a single interspecies spillover from wild birds followed by sustained cow-to-cow transmission for several months prior to initial identification.^3^

Three human cases of H5N1 HPAI have been associated with the dairy cattle outbreak in the United States, the first occurring in Texas and the second two in Michigan. All individuals were dairy workers and had presumed exposure to infected cattle, indicating a new risk factor for infection in humans.^4-6^

To date, there have been no reported incidents of H5N1 HPAI in Canadian cattle. The Canadian Food Inspection Agency (CFIA) in collaboration with the Public Health Agency of Canada (PHAC) and Health Canada conducted testing of 303 commercially available milk products across Canada, including 75 from Ontario, all of which were negative for influenza A viral RNA.^7^

Ontario has the second largest number of dairy farms and dairy cattle in Canada^8^ and borders several US states with active outbreaks, including Michigan. Due to unanticipated H5N1 HPAI outbreaks in cattle and the possibility of cryptic transmission among cows, we undertook a pilot study to assess the presence of influenza A vRNA in commercially available pasteurized milk products in the province of Ontario.

## Methods

This study included pasteurized cow’s milk purchased from commercial vendors in the province of Ontario. Cream products (5% fat content or higher), flavoured (e.g. chocolate) milk, cheese and other dairy products were excluded. Ultra-high-temperature (UHT) and microfiltered milk were accepted. Samples were sourced from microbiology laboratory and related research staff in Ontario and transported in clean or sterile containers, then aliquoted and frozen at -80°C pending testing.

Samples were thawed and vortexed, then diluted 1:4 (vol/vol) in sterile phosphate buffered saline (PBS) solution^9^. RNA extraction was conducted via Nuclisens Easymag following generic protocol 2.0.1 with 140uL input volume and 40uL elution volume. Armored RNA enterovirus (arm-ent) (Asuragen; https://www.asuragen.com) spiking was used as an internal extraction control. Samples were tested by quantitative polymerase chain reaction (qPCR) on the Quantstudio 3 platform with the Luna Universal One-Step RT-qPCR Kit (NEB #E3005). Samples were tested in duplicate with 5uL of RNA template and the following cycling conditions with fast qPCR conditions: hold stage: 55°C for 10 min, 95°C for 1 min followed by 44 amplification cycles of 95°C for 10 sec and 60°C for 30 sec. PCR assay multiplex included US Centers for Disease Control and Prevention universal influenza A matrix gene primer and probe set^10^ (below) and arm-ent primer and probe set.

CDC_FluA (probe): 5’ d FAM-TGCAGTCCTCGCTCACTGGGCACG-BHQ-1 3’

CDC_FluA_Forward: 5’ d GACCRATCCTGTCACCTCTGAC 3’

CDC_FluA_ Reverse: 5’ d AGGGCATTYTGGACAAAKCGTCTA 3’

Limit of detection (LOD) of the assay for vRNA in milk samples was determined by spiking 2% milk with influenza A H3N2 and pasteurizing at 72°C for no less than 16 seconds as is most common in Ontario^11^. The limit of detection (LoD) for the assay with the above protocol is a final viral concentration of 10^2^ pfu/mL. The following processes were tested during assay optimization and provided no significant improvements to LoD: centrifuging milk samples prior to extraction at 4°C and 6,000xg for 5 minutes, filtering milk samples prior to extraction with 0.45um filter, increasing extraction input volume to 600uL or 200uL, adding additional wash steps to the extraction (Easymag specific B 2.0.1 protocol), conducting a magnetic bead clean-up on RNA template prior to PCR, and decreasing RNA template volume to 3uL, 2.5uL, 2uL, or 1uL.

## Results

One hundred and seventeen commercially available cow’s milk samples were received by Sunnybrook Research Institute (SRI) from 17 different municipalities across Ontario (Figure 1, Table 1). Overall, 28 brands were sampled: 6 skim milk, 24 1%, 56 2%, 31 whole milk, with best before dates ranging from April 15, 2024 to August 28, 2024. Two samples were provided as controls from milk that had been frozen at -20C for approximately 18 months. All samples tested negative for influenza A target by qPCR with internal extraction control (arm-ent) target cycle threshold values (Cts) ranging from 31 to 40.

**Figure 1.**
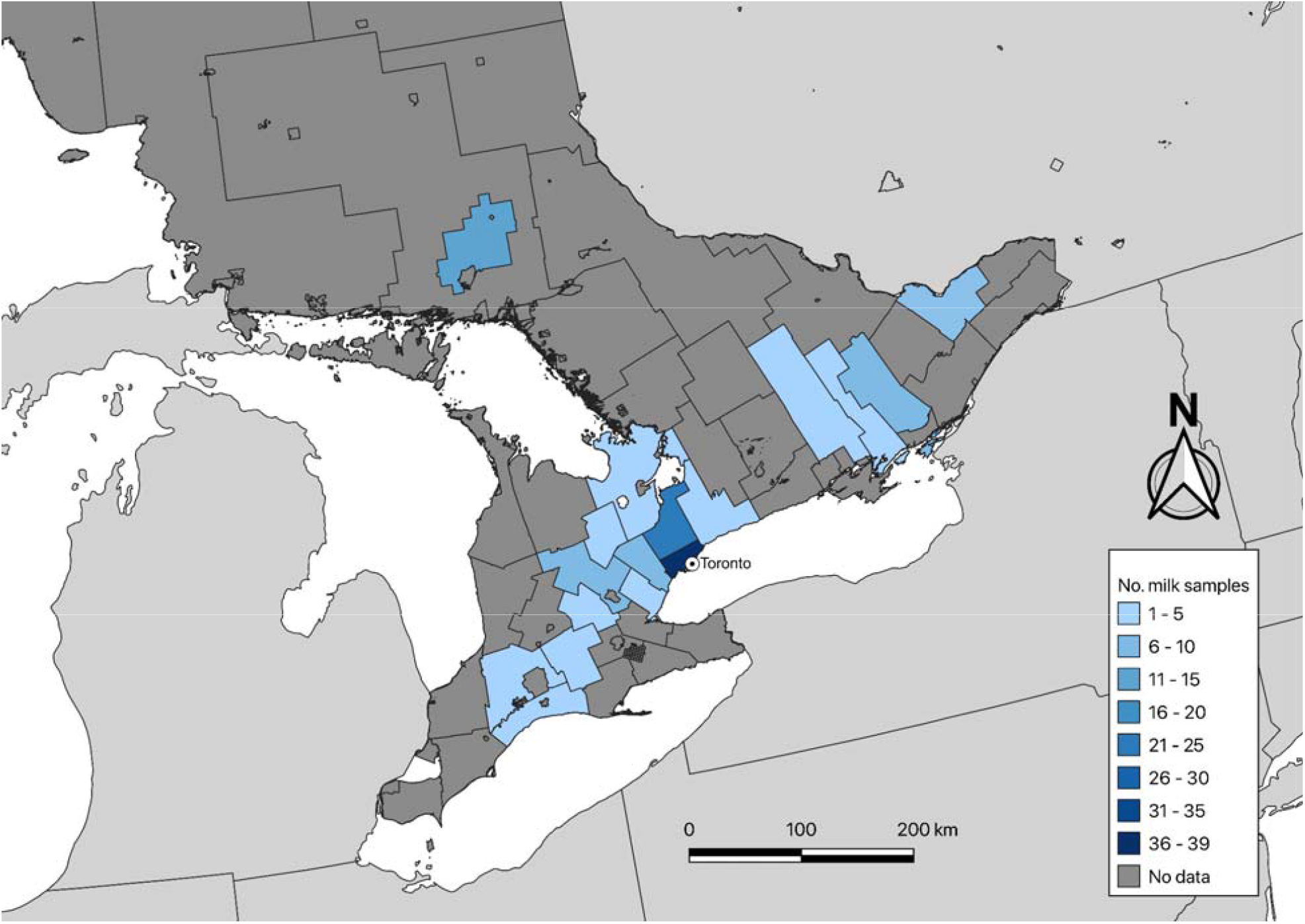
Locations of purchase of commercially available milk samples. This map of the province of Ontario, Canada, indicates the upper-tier and single-tier (cities of Toronto, Ottawa, and Greater Sudbury are single tier) municipalities where the commercial milk samples for this study were purchased. Colour indicates the number of samples purchased at each municipality. The city of Toronto is labeled.

## Conclusion

We did not detect influenza A viral RNA in any of the commercially available Ontario milk products included in this study. The upper 95% confidence limit estimate from a sample set of this size suggests that fewer than 4% of milk samples would potentially contain viral RNA, significantly less than that detected in US sampling. However, our sample does not include milk from all regions of the province, and the sample size does not rule out the possibility of H5N1 HPAI circulating in Ontario cattle that may have been acquired after our sampling period.

## Data Availability

All data produced in the present study are available upon reasonable request to the authors

## Acknowledgements

The authors thank the microbiology laboratory and research staff who provided the samples for testing as well as Dr. Andrew Bowman from the Ohio State University and Dr. Davor Ojkic from the Animal Health Laboratory (Guelph) for providing guidance on the design and optimization of methods.

